# Recruiting Busy Clinicians through Asynchronous Online Focus Groups: Recruitment Evidence from Two Projects

**DOI:** 10.1101/2025.09.19.25336092

**Authors:** Mario Venegas, Richard SoRelle, Amari S. Anderson, Traber D. Giardina, Jennifer Freytag, Sylvia J. Hysong

## Abstract

**Objective:** Due to an increase in healthcare quality research and QI projects, there is a high demand for clinicians’ time to serve as research participants. Traditional focus group methods are time demanding, posing a barrier to collect data from busy clinicians. We show that asynchronous online focus groups (AOFGs) are a viable method for increasing participation from busy clinicians.

**Materials and Methods:** Recruitment metrics were computed by counting the number of enrollees and sites of provenance during the period when traditional focus groups and AOFGs were offered, respectively. Then, using Microsoft Teams, private channels served as forums for primary care staff at Veterans Health Administration to share their experiences as part of two healthcare projects.

**Results:** 82 primary care staff from 17 VA sites individually participated. Pivoting to AOFGs helped increase acceptance rates by 429% and 116% in each respective project. Offering AOFGs helped decrease non-response rates across both projects by 35% and 16.34% on both projects. Notably, Care Managers (RNs) were the most well-represented, constituting nearly 40% of participants on each project.

**Discussion:** This paper explores metrics that can be tracked and measured for future research. AOFGs replace the coordination work of gathering participants in one place in real time with the protracted coordination work of keeping participants engaged. AOFGs thus require consistent communication between recruiters and participants as well as active moderation to sustain engagement.

**Conclusions:** By providing a flexible alternative that accommodates the unpredictable demands clinicians face in primary care, AOFGs help address the barrier of time demand and increase participation in quality improvement and healthcare research projects.

## BACKGROUND AND SIGNIFICANCE

Due to an increase in Quality Improvement (QI) projects and in healthcare quality research, there is an increased demand for clinician time to serve as research participants.^1, 2^ Further, the growing requests for participation in research -- particularly for healthcare professionals who were affected by emergencies like COVID-19 -- contributes to research fatigue, i.e. the exhaustion from individuals fielding multiple requests to participate in research.^3, 4^ Indeed, when the benefits of engaging in research cannot offset the time costs, research fatigue can occur among healthcare professionals.^3^ Qualitative data collection methods, by design, are obtrusive as they often demand time away from job duties for healthcare workers.^3^

Among qualitative methods, traditional focus groups are time consuming and require significant schedule cooordination^5, 6^, posing a recruitment barrier for busy clinicians who often attend to unpredictable emergencies and demands from work.^7, 8^ The fast-paced environment of primary care, which includes the intensive labor of providing care, filling out administrative paperwork, and scrubbing records creates logistical difficulties for participants to commit an hour or more of their time as research participants.^1, 2, 9^ Thus, time-adaptable data collection methods are needed to collect qualitative data from this high-demand and busy population. Asynchronous Online Focus Groups (AOFGs), provide a flexible option to accommodate the fast-paced and unpredictable environment of primary care.

Researchers began using AOFGs in the 1990s with the rise of the internet.^10^ Starting out in chat rooms and discussion boards, researchers adapted AOFGs to social media applications such as Facebook, WhatsApp, and Microsoft Teams^11, 12^. As focus group methods went virtual in real-time online meetings, AOFGs, despite happening in channels that are self-paced, still retain the core feature of “gathering data through group interactions regarding a phenomenon determined by the researcher”^7^. AOFGs still collect shared understandings and views about a defined topic, and the flow of the conversation is shaped by a moderator, with participants and moderators replying at their own time. A growing body of literature about AOFGs showcases applications across multiple contexts and populations such as healthcare patients, healthcare professionals, data collection during COVID-19’s social distancing policies, among adolescents and university students^12-15^. However, although AOFGs are suitable in situations where time and resources are constrained^10, 16^ and can accommodate routine and unpredictable demands in the workplace, we know little about how this method impacts participant recruitment.

### objective

To address this gap in the literature, we explore how AOFGs impact participant recruitment. We draw on recruitment data from two projects, and supplement with research team recruiter reflections, to determine the extent to which asynchronous online focus groups (AOFGs) are a viable method for increasing participation from busy clinicians in research and QI.

## MATERIALS AND METHODS

We show how our application of AOFG methods impacted participant recruitment as part of two larger separate studies: a traditional research study and a healthcare operations evaluation. Both took place in primary care in the Veterans Health Administration (VHA).

### project context

In both projects, we collected qualitative data from primary care clinicians about their experiences with how preventive health screenings for cancer and mental health were conducted at their respective VA Medical Centers. The research project examined the role of team coordination in the context of maintaining preventive care during the COVID-19 Pandemic. The other project was an operations evaluation of local workflow adaptations by VHA Primary Care clinics. Each project involved collecting qualitative data from facility leadership, front-line clinicians, and —in the research study—from Veterans. This paper focuses on the data collection from front-line clinicians, which were originally planned as synchronous focus groups via Microsoft Teams. We asked clinicians to share their challenges with conducting routine screenings, and to share how from their perspective the practice of preventive health screening had changed during pre-established study observation periods—both of which covered the COVID-19 Pandemic. Due to practical challenges such as time and unpredictable demands that led participants to cancel the scheduled synchronous focus groups, we pivoted to AOFGs to accommodate these factors.

### participant recruitment

Research coordinators, who were part of the research team, functioned as recruiters for both projects. They contacted participants via email first, then through Teams private message if there was no email response within 1-2 days. If interested, recruiters consented participants and informed them of the process for engaging in AOFGs. Recruiters tracked participant data and individual contact attempts in a custom-designed Microsoft Access database. Data on each contact included the date and time of the contact, the person making the contact attempt, the type of contact, the contact result, and an open-ended notes field for recording any additional information not captured in structured form fields. The “contact result” field allowed recruiters to enter “declined”, “ineligible”, “accepted”, “scheduled”, “rescheduled”, “completed”, and “no response”.

### recruitment data measures

Although the main project drew on qualitative asynchronous focus group data, the recruitment database in Microsoft Access captured quantitative metrics for participant recruitment, retention, and attrition. Table 1 lists the main metrics we used to capture clinician recruitment throughout both projects’ timelines, both before and after AOFGs. Recruitment success (enrollment) was calculated by running queries counting the numbers of invitees with contact results in each category, typically by site. Through these queries, we measured enrollment during the period when we offered traditional focus groups via Teams video call -- 1/22/2024 for the evaluation, and 8/16/2023 for the research study-- and compared it to the period when we offered AOFGs via Teams channels-- 4/22/2024. Lastly, we collected recruiter testimonials about their experience contacting clinicians to help supplement the database metrics and to provide contextual dimensions about their experience.

**Table 1:** Metrics Quantifying Recruitment Contacts for Asynchronous Online Focus Groups Across Both Projects.

| Metric | Definition | Service Directed Research Study |  |  |  | Operations Evaluation Project |  |  |  |
| --- | --- | --- | --- | --- | --- | --- | --- | --- | --- |
|  |  | Mean<br>or % | SD | Min-Max | n | Mean<br>or % | SD | Min-Max | n |
| Contacting Participants | *Number of contact attempts to a potential participant prior to receiving an initial response* | 3.0 | 2.2 | 1-19 | 419 | 2.3 | 1.8 | 0-11 | 153 |
| Initial Response Time | *Cycle time in calendar days from initial contact attempt to initial response.* | 154.4 | 128.8 | 0-471 | 419 | 89.2 | 87.1 | 0-279 | 153 |
| No Response | *Percent of potential participants who did not respond to contact attempts* | 49% | -- | -- | 744 | 77.5% | -- | -- | 564 |
| Declined | *Percent of potential participants who declined (i.e. those who responded to recruiters and were not lost to contact or deemed ineligible)* | 22.6% | -- | -- | 744 | 11.9% | -- | -- | 564 |
| Ineligible | *Percent of potential participants who were deemed ineligible to participate either by their own choice or by recruiter criteria.* | 20.6% | -- | -- | 744 | 4.1% | -- | -- | 564 |
| Accepted | *Percent of potential participants who agreed to be part of the AOFG.* | 7.9% | -- | -- | 744 | 6.6% | -- | -- | 564 |
| Completed AOFG | *Number of contacts stating that participant completed their participation in the AOFG.* | 80.3% | -- | -- | 56 | 94.6% | -- | -- | 56 |
| Left AOFG | *Participants leaving the AOFG, either by explicitly telling the recruiter, no longer responding when AOFG started, or by leaving the Teams channel with no communication.* | 19.6% | -- | -- | 56 | 5.4% | -- | -- | 56 |

### AOFG platform and procedures

Using Microsoft Teams private channels, we created a separate AOFG channel for each participating VA site and added participants who consented to participating to each channel. Microsoft Teams channels allowed for privacy such that participants could only access channels to which they had been added by the study team. Over the course of three weeks, we engaged participants via channel posts eliciting comments. The first week focused on challenges with preventive health screenings to help foster conversation with participants and familiarize them with the routine of daily posts and reminders. Through this process, the moderator was able to ask follow-up questions and sustain the conversation. On the second week, we shifted to eliciting comments on four screening process maps: breast cancer, colorectal cancer, depression, and PTSD. The moderator posted on the channel and attached an image of the process flow for the screenings, asking participants to share if the process looked accurate to their practice or if there were corrections to be made. This process lasted between seven and twelve days, depending on participant activity. During the final week, we completed the process map discussions and shifted to the closing question for how the practice of primary care changed for them since COVID. Depending on activity from participants, the moderator was able to ask further follow-up questions to probe deeper into participants’ experiences. Lastly, as a closing message, the moderator thanked all participants for their time and provided links to the previous questions in case participants wanted to post any further responses or if participants missed some questions.

As a supplement and a means to manage comments on process maps, we utilized Microsoft Teams’ built-in Whiteboard application. This interactive feature allowed the research team to post process map images and post comments in the form of virtual sticky notes. The moderator would copy the responses from participants on the channel and post their comments as sticky notes in the respective process map to which participants referred. We initially used the Whiteboard as part of the data collection process, asking participants to post their sticky notes and we included instructional videos and posted Q&As with repeated information on how to navigate this app. Due to the learning curve with navigating Whiteboard, we simplified posting process map images and elicit responses on channel posts instead.

### data analysis

Analysis consisted of descriptive statistics measuring recruitment metrics before and after we pivoted to AOFGs. These statistics measured rates of acceptance, rate of completion of AOFGs among those who accepted, rates of participants declining, and rates of no response from participants. In addition to descriptive statistics, we also included recruiter testimonials to provide experiential data on the recruitment process both before and after we shifted to AOFGs.

## RESULTS

### participant recruitment before and after AOFGs

As shown in Table 1, the acceptance rate for both projects was 7.9% for the research study and 6.6% for the operations evaluation project. Additionally, those who agreed to participate showed a high completion rate for both projects. The research study had an 80.3% completion rate while the operations evaluation had nearly 95% of participants complete their participation. Across both projects, the initial response time was relatively large, with a wide deviation in response time. However, the operations project had a relatively shorter response time (average 87.1 days) than the research study (average 154.4 days). Although the research study had a nearly 50% non-response rate, the operations project had a 77.5% non-response rate from participants. Still, the rate of declined contacts was higher for the research study, 1.90 times as much (22.6%), as it was for the operations project (11.9%). Notably, the rate of ineligible participants was nearly 5 times higher in the research study (20.6%) than it was in the operations project (4.1%).

Table 2 provides a breakdown of clinicians who participated in the AOFGs. Participants included Medical Support Assistants (MSA), Primary Care Providers (PCP), Care Managers (RNs) and Clinical Associates (LPNs). In both projects, nearly the same number of clinicians participated in the AOFGs-43 in the research study, 39 in the operations project. Notably, Care Managers (RNs) were the most represented across both projects, constituting nearly 40% of participants in each sample. Nearly twice as many PCPs participated in the research study (25.6%) as they did in the operations project (12.8%). Further, Administrative Associates (MSAs) constituted nearly 30% of the sample for the operations project while they were only 19% of the research study. Lastly, the operations project ended up with care team extenders, including one psychologist and one social worker who were able to provide their own perspectives on maintaining preventive care.

**Table 2:** Primary Care Team Role Distribution of AOFG Participants.

| PACT Role | Service Directed Research Study |  | Operations Evaluation Project |  |
| --- | --- | --- | --- | --- |
|  | N | % | N | % |
| Administrative Associate | 8 | 18.6 | 11 | 28.2 |
| Care Manager | 17 | 39.5 | 15 | 38.5 |
| Clinical Associate | 6 | 14.0 | 6 | 15.4 |
| Primary Care Provider | 11 | 25.6 | 5 | 12.8 |
| Psychologist | 0 | 0 | 1 | 2.6 |
| Social Worker | 0 | 0 | 1 | 2.6 |
| Other | 1 | 2.3 | 0 | 0 |
| Total | 43 | 100.0 | 39 | 100.0 |

Prior to pivoting to AOFGs, recruitment consisted of contacting clinicians to schedule an online meeting in real time together. When stratified by before vs. after AOFGs, the number of participants recruited differed with approximately twice as many participants contacted before AOFGs in both projects (See Figure 1). With these differences in sample sizes, we compared recruitment metrics from before and after pivoting to AOFGs, breaking down the overall metrics in Table 1. We captured those who left the AOFG but not the participants who accepted the traditional online focus group but did not commit to join the conversation.

**Figure 1:**
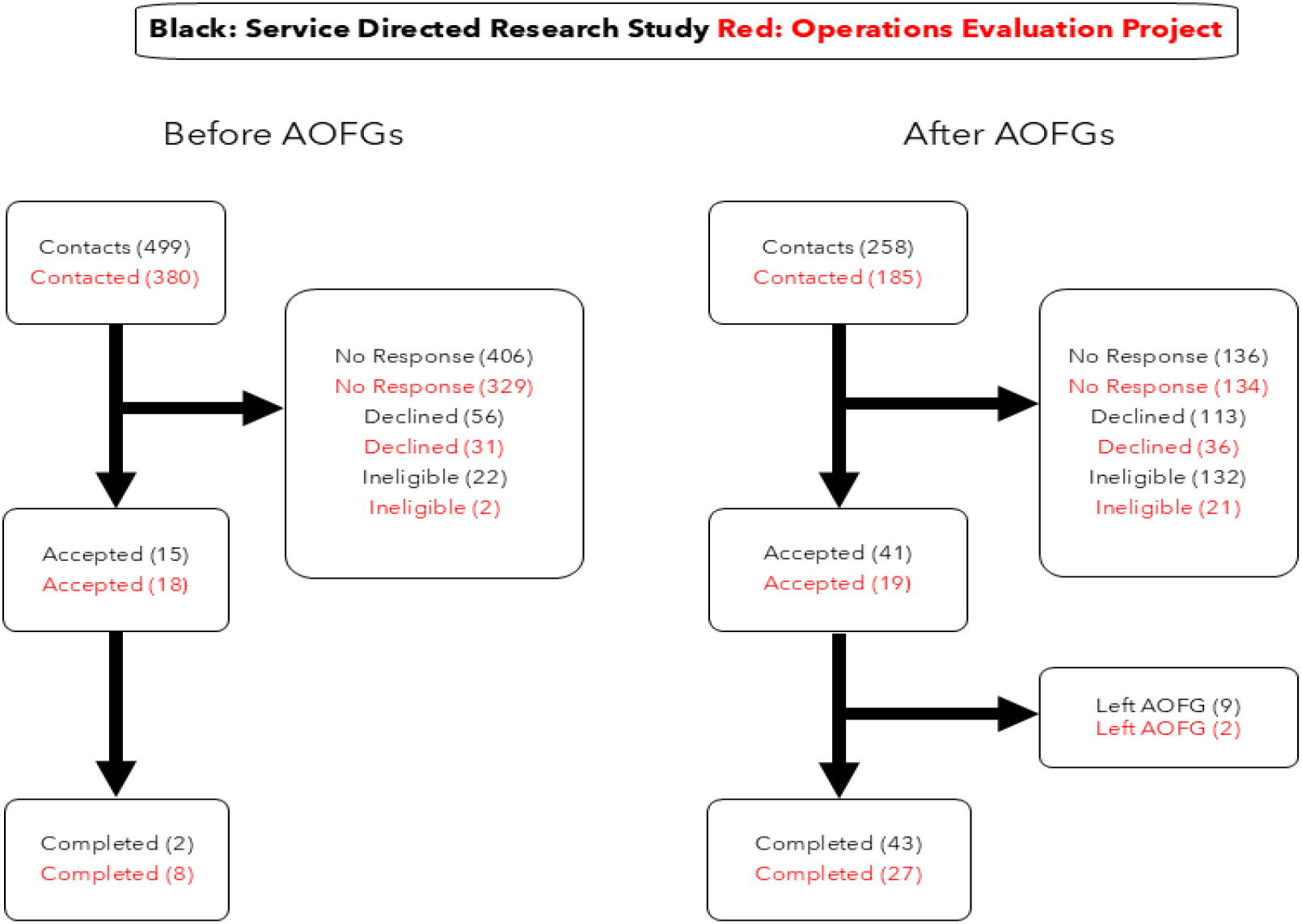
Recruitment Contacts Before and After AOFGs.

For the service-directed research study, recruitment initially relied on real-time focus group video calls, generating an acceptance rate of 3.01%. The introduction of asynchronous focus groups (AOFGs) increased the acceptance rate by 429% to 16% (See Table 3). Similarly, the completion rate improved by 4058%, rising from 0.40% to 17% after the implementation of AOFGs. The proportion of participants who did not respond to recruitment attempts also decreased by 35%, from 81.36% to 53%. However, the rate of participants who declined participation in the study increased by 290%, from 11.22% to 44%.

**Table 3:** Recruitment Metric Changes Before and After AOFGs.

| Metric | Service Directed Research Study |  |  |  |  | Operations Evaluation Project |  |  |  |  |
| --- | --- | --- | --- | --- | --- | --- | --- | --- | --- | --- |
|  | Before | After | % before |  | %change | Before | After | % before |  | % change<sup>1</sup> |
|  | AOFG | AOFG | before | %after |  | AOFG | AOFG | before | % after |  |
| Contact | 499 | 258 |  |  |  | 380 | 185 |  |  |  |
| No Response | 406 | 136 | 81.36% | 53% | -35% | 329 | 134 | 87% | 72.43% | -16.34% |
| Declined | 56 | 113 | 11.22% | 44% | 290% | 31 | 36 | 8% | 19.46% | 138.54% |
| Ineligible | 22 | 132 | 4.41% | 51% | 1060% | 2 | 21 | 1% | 11.35% | 2056.76% |
| Accepted | 15 | 41 | 3.01% | 16% | 429% | 18 | 19 | 5% | 10.27% | 116.82% |
| Left AOFG |  | 9 | 0.00% | 3% |  |  | 2 | 0% | 1.08% |  |
| Completed | 2 | 43 | 0.40% | 17% | 4058% | 8 | 27 | 2% | 14.59% | 593.24% |

<sup>1</sup>Ratio of the difference between the percentage before and percentage after AOFGs to capture relative change.

Similarly, for the operations evaluation project, the acceptance rate increased by 116.82%, going from 5% before AOFGs to 10.27% after AOFGs. The completion rate improved by 593.24%, increasing from 2% to 14.59%. Additionally, the number of participants who did not respond to any recruitment contacts decreased by 16.34%, from 87% to 72.43%. However, the proportion of participants who actively declined participation increased by 138.54%, from 8% to 19.46%. Lastly, by offering a flexible alternative that accommodates the busy schedules of clinicians, the number of participants who completed the focus groups increased by 593.24%, going from a 2% completion to 14.59% completion. Although sample sizes before and after AOFGs differed, both projects pivoted to AOFGs at similar timelines. As shown in Figure 2, AOFGs were introduced at comparable intervals for the operations project, allowing for a consistent timeline of recruitment and allowing for a reliable comparison across the two studies.

**Figure 2:**
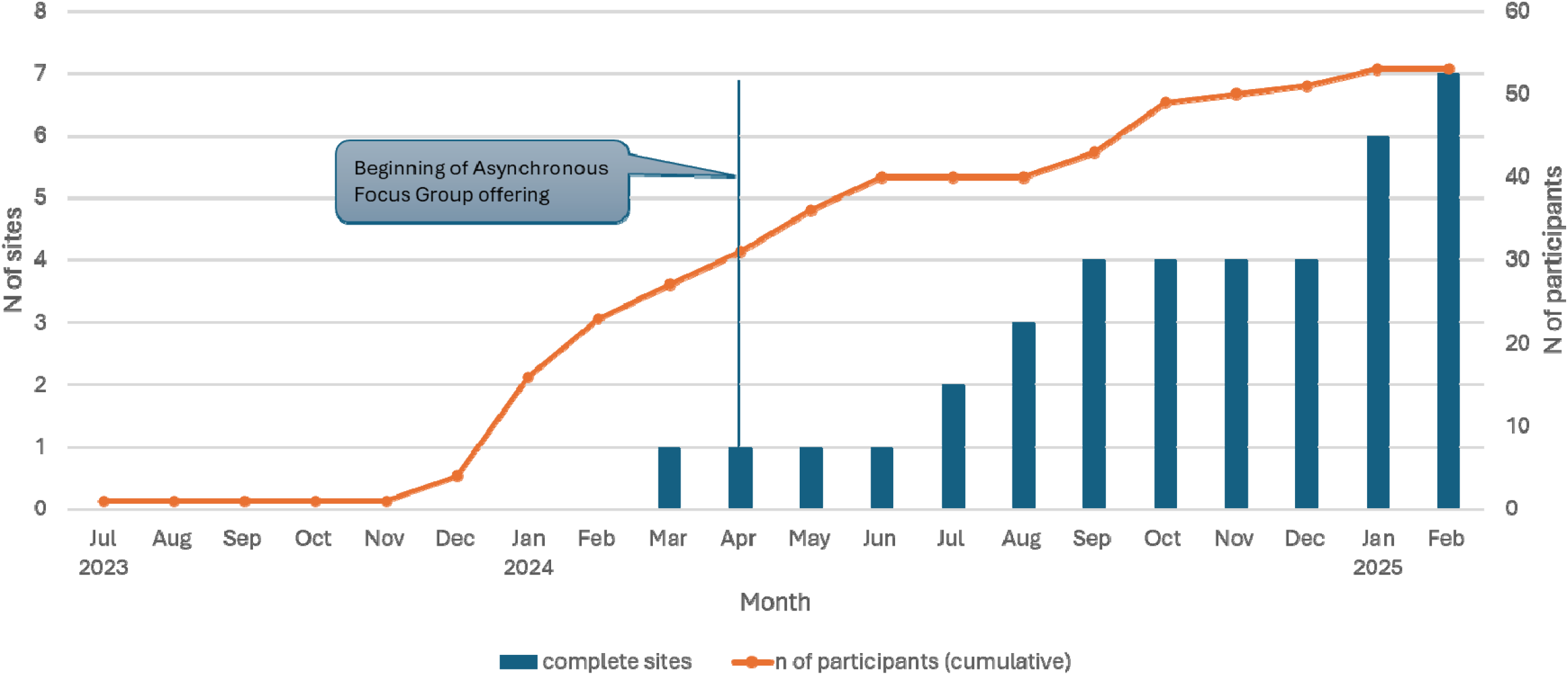
Number of VHA Sites and Participants Recruited Over Life of Healthcare Operations Project.

### recruiter reflections

We gathered insights from members of the research team (n=4) who were directly involved in participant outreach. These recruiters played a crucial role in engagement efforts, and their reflections helped illuminate the nuances behind recruitment numbers. To preserve their anonymity, pseudonyms are used below.

Recruiters reported notable differences in participant responsiveness depending on the format of the focus group offered. During the initial phase of the project, when synchronous video focus groups were the primary method, recruiters experienced substantial rejection. Valery, one of the recruiters, described the difficulty in scheduling live sessions with clinicians:

> *“When we were following the original plans to set up a Microsoft Teams call focus group, many clinicians would often say no simply because they did not have time in their schedule to sit down for an hour and complete a focus group. So in the beginning of my recruitment phase, it did have a lot of rejection from the clinicians*.*”*

However, following the implementations of AOFGs, recruiters observed a shift in responsiveness. Valery noted, “*but once we change[d] the format to completely asynchronous, I was able to go back and recruit some clinicians who had previously told me no*.” She continued,

> *“I started getting more yesses immediately. I’m not sure what the exact date was that we got greenlit to start saying the focus group would be asynch [,] but once I sent out my first recruitment message including that detail [,] I got 2 yesses that day, and the rest trickled in within the next couple of weeks. For my sites that I was able to immediately start recruitment with the asynch info I was able to recruit at least 5-8 participants within a month of my first contact, the quickest site I had only spent 2 weeks recruiting!”*

Claire, another recruiter, echoed these experiences, explaining that clinicians responded more positively to AOFGs because they offered flexibility: *“they were more favorable of the option [AOFG] since they could personalize it to their own schedule*.*”* These firsthand reflections underscore the importance of format flexibility in recruitment strategies, particularly when engaging clinicians who have limited and unpredictable workloads. Recruiter testimonies not only support the observed quantitative trends but very importantly, also highlight the notable change in participant response when offered the asynchronous option.

## DISCUSSION

We sought to show AOFGs were a viable method to increase participation from busy clinicians in QI and research projects. The relatively high percentages of RNs and MSAs participating underscores the ability for AOFGs to capture these busy populations who handle the fast-paced demands of primary care. In illustrating recruitment data, our findings build on the AOFG literature in three ways:

1. As shown in Figure 1 and Table 3, pivoting to AOFGs helped increase acceptance rates and decrease the non-response rate. Testimonies from team recruiters illustrated AOFGs help accommodate participant’s schedules. Given the population of VHA clinicians whose workload has increased since COVID, these findings are consistent with studies which assert AOFGs are flexible to individuals’ schedules and when time is constrained.^10, 13, 14^
2. Although literature on AOFGs has explored the implementation of AOFGs across various online platforms,^11, 17^ how the method worked across healthcare contexts,^13^ and gleaned lessons learned in implementing AOFGs,^14^ there is little research on the impact of this method on participant recruitment. Our paper illuminates how this method increased participation by way of higher acceptance rates after pivoting to AOFGs for both projects. Moreover, our paper also showcases metrics that can be tracked and measured using database software. The data can further be analyzed for associations beyond descriptive statistics, and this paper helped build on metrics that researchers can use to track recruitment efforts in healthcare research and to measure how AOFGs are received by populations in the healthcare sector.
3. Recruitment findings illustrate lack of time does not equate to lack of interest, despite how busy clinicians have become. By providing a flexible alternative, adapting to demands clinicians face, AOFGs help address one barrier driving research fatigue: time demand.^3, 4^ However, in pivoting to AOFGs, there is a trade-off in coordination work. AOFGs replace the coordination work of gathering participants in one place in real time with the protracted coordination work of keeping participants engaged during the entire three-week period. Thus, AOFG engagement requires active communication between recruiters and participants, and as shown in past studies, active moderation and participation from the facilitator.^12, 14^ Despite the increase in participant recruitment for research and QI projects using AOFGs, we note key limitations.

### limitations

Although we captured recruitment data, we did not capture data from participants on their experiences with the AOFG format. Due to the self-paced and online mediated nature of AOFGs, we cannot see participant reactions or capture live responses in real time. Some ways to capture cues left out by the online format has been through documenting emojis, punctuation, tagging, and misspelling^18^. Nevertheless, our paper presents a recruitment perspective on AOFGs, and future studies should capture participant views on this method and whether they prefer AOFGs and further explore under what conditions this method is viable.

Similarly, we could not compare focus group data from traditional synchronous focus groups and asynchronous focus groups and thus we cannot compare data quality. However, our paper provided evidence that pivoting to AOFGs in the context of busy clinicians with unpredictable work schedules increased recruitment. In addressing the question of data quality, Woodyatt and colleagues showed that sensitive topics were discussed more candidly in online synchronous focus groups compared to in-person focus groups.^19^ For AOFGs, Frey and Bloch note that researchers can summarize ongoing conversations to ensure they interpret participants’ responses to mitigate the lack of quality responses.^12^ Still, more research is needed comparing content quality between synchronous and asynchronous focus groups.

Lastly, due to the types of questions we set out to investigate in both projects, the AOFGs were designed to include primary care personnel from Medical Support Assistants to Primary Care Providers. Thus, we could not account for status hierarchies and power relations among primary care personnel that may shape how participants respond on a private Teams channel. Future healthcare projects can account for these socio-cultural factors by designing AOFGs for each type of personnel: nurses, medical support assistants, etc. Despite these limitations, our findings show that AOFGs provide a flexible option that adapts to the unpredictable demands in the primary care workplace.

## CONCLUSION

Using recruitment data and recruiter testimonials, AOFGs were a viable method to increase participation from busy clinicians in QI and research projects. Thus, these results illustrate that lack of time does not equate to lack of interest for a population that has become increasingly busy since the COVID-19 Pandemic. AOFGs are suitable for circumstances where time is precarious. However, there is still need for more research on how AOFGs can obtain and sustain quality and richness of qualitative responses, what best practices can help in obtaining rich qualitative data, and what are drawbacks to using this method from participants who may be overflooded with messages. Nevertheless, to echo Frey and Bloch, we are not arguing for replacing traditional real-time focus groups^12^. Rather, pivoting to this method helped us overcome the issue of time-demand from participants whose time and resources were constrained due to their job duties. Capturing and providing recruitment metrics provides departure points for future quantitative analyses that can examine recruitment strategies, frequency of outreach, among other variables and for which researchers can examine their associations with acceptance rates among a population burdened with job and research demands.

## Acknowledgements

I would like to acknowledge the contributions of Dr. Jennifer Sloane, Dr. Traber Giardina, Dr. Jennifer Freytag, and Jessica Castillo for their edits, revisions, and comments on the working draft. Lastly, I would like to acknowledge the HERMES team for their support, comments, and suggestions in preparing this manuscript.

## Declaration Statement

The authors declare that they have no known competing financial interests or personal relationships that could have appeared to influence the work reported in this paper.

## Data Availability Statement

The data that support the findings of this study are available upon written request from the corresponding author. The data are not publicly available, per VA privacy policies.

